# Immune Cell Profiles and Cytokine Levels of Patients with Interstitial Cystitis/Bladder Pain Syndrome

**DOI:** 10.1101/2020.12.17.20248414

**Authors:** Robert M. Moldwin, Vishaan Nursey, Oksana Yaskiv, Siddhartha Dalvi, Michael Funaro, William DeGouveia, Marina Ruzimovsky, Horacio R. Rilo, Edmund J. Miller, Souhel Najjar, Inna Tabansky, Joel N.H. Stern

**Affiliations:** The Smith Institute for Urology, Northwell Health, 450 Lakeville Road New Hyde Park, NY; Department of Urology, Donald and Barbara Zucker School of Medicine at Hofstra/Northwell, Hempstead, New York, USA; Department of Neurology, Donald and Barbara Zucker School of Medicine at Hofstra/Northwell, Hempstead, New York, USA; Institute of Molecular Medicine, The Feinstein Institutes for Medical Research, Manhasset, NY, USA; Department of Pathology, Donald and Barbara Zucker School of Medicine at Hofstra/Northwell, Hempstead, New York, USA; Department of Surgery, Donald and Barbara Zucker School of Medicine at Hofstra/Northwell, Hempstead, New York, USA; RDS2 Solutions. Stony Brook, NY, USA; Department of Neurobiology and Behavior, The Rockefeller University, New York, NY, USA; Department of Neurology, Lenox Hill Hospital, New York, NY, USA; Renaissance School of Medicine at Stony Brook University, Stony Brook, NY, USA

**Author notes:** To whom correspondence should be addressed Correspondence: Joel N.H. Stern (jstern5@northwell or) or Robert M. Moldwin, The Smith Institute for Urology, 450 Lakeville Road, New Hyde Park, NY, 11042. authors contributed equally (Robert M. Moldwin, Vishaan Nursey).

**Keywords:** Plasma Cells, B-cells, Interleukin-6, Histology, Biopsy, Hunner’s Lesions, Urine

## Abstract

**Aims:** To quantify the number of immune cells in the bladder urothelium and concentrations of urinary cytokines in patients with Interstitial Cystitis/Bladder Pain Syndrome (IC/BPS). To identify differences in these measures in IC/BPS patients with Hunner’s lesions (IC/BPS-HL) and without Hunner’s lesions (IC/BPS-NHL).

**Methods:** Bladder tissue biopsies were obtained from 48 patients with IC/BPS-HL and unaffected controls (UC) and stained with antibodies for various immune cell markers such as CD138, CD20 and CD56. Levels of cytokines (Interferon (IFN)-γ, Interleukin (IL)-1β, IL-2, IL- 4, IL-6, IL-8, IL12P70, IL-13, and TNF-α) were measured from normalized urine obtained from 18 IC/BPS-HL, 18 IC/BPS-NHL, and 4 UC.

**Results:** Numbers of CD138+ plasma cells, CD20+ B cells, and CD3+ T cells were significantly increased (50 fold, 30 fold, and an almost 3 fold increase, respectively; p-values: 1.34E-06, 3.26E-04, and 2.52E-6) in the bladders of IC/BPS-HL patients compared to UC. Patients with IC/BPS-HL had significantly elevated urinary levels of IL-6 (p=0.0028) and TNF-α (p=0.009) compared to patients with IC/BPS-NHL and UC. In contrast, IL-12p70 levels were significantly higher in the patients with IC/BPS-NHL than in HL patients (p=0.033). No significant difference in IL-12p70 levels were observed between IC/BPS-HL and UC.

**Conclusion:** Different cytokines were elevated in the urine of IC/BPS patients with and without HL, suggesting differences in underlying disease processes. Elevated levels of CD138+, CD20+, and CD3+ cells in HL indicate B and T-cell involvement in lesion formation. Determining which cytokines and immunological pathways are present in IC/BPS-HL could elucidate the disease mechanism.

## Introduction

Interstitial Cystitis/Bladder Pain Syndrome (IC/BPS) affects millions of individuals within the US.^1^ It is defined by bladder pain/discomfort accompanied by urgent voiding symptoms in the absence of other identifiable pathology. Most afflicted individuals have some portion of their pain derived from extravesical sources. IC/BPS is often difficult to diagnosis because symptoms are similar to other disorders, such as overactive bladder, vulvodynia, endometriosis, and prostatitis.^2^ Identification of targets for diagnostic tests and treatments that consistently and accurately predict disease status and/or provide insights into disease mechanisms is an ongoing challenge.^3^

While most patients with IC/BPS show no lesions or ulcers within their bladders, (IC/BPS-NHL), A relatively small percentage of patients with IC/BPS present with focal regions of gross bladder inflammation termed Hunner’s lesions (HL). These lesions are typically associated with small vessels radiating toward a central scar and a fibrinous exudate (**Figure 1**).^4^ It is unclear whether IC/BPS with Hunner’s Lesions (IC/BPS-HL) represents a point on the spectrum of IC/BPS, or a separate disease process altogether. Determining which cytokines and immunological pathways are present in HL could help identify whether it is part of the spectrum of IC/BPS-NHL or a unique disease.

**Figure 1:**
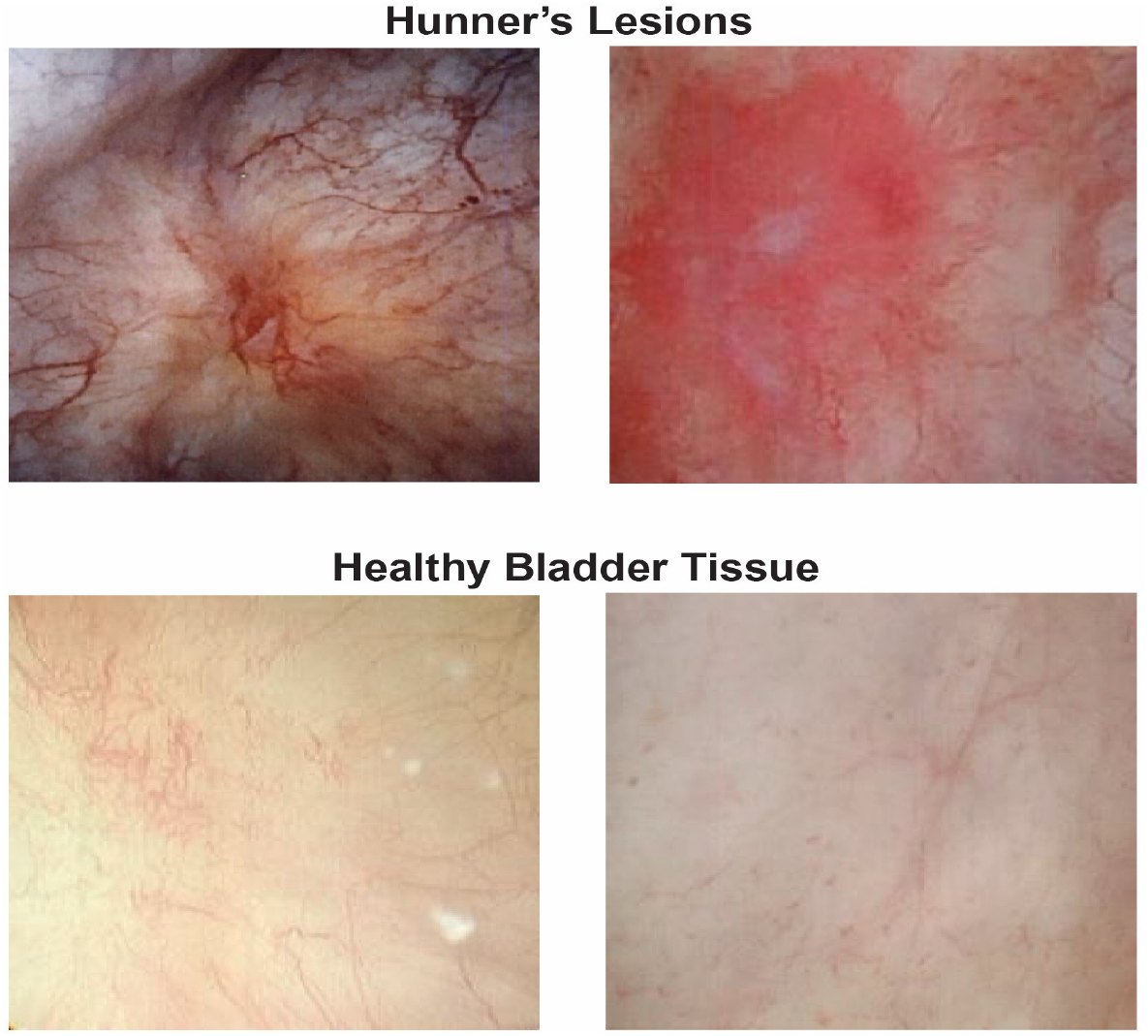
Cystoscopy images of Hunner’s Lesions (HL) from 2 different patients with Interstitial Cystitis/Bladder Pain Syndrome with Hunner’s Lesions (IC/BPS-HL) compared to healthy bladder tissue. HLs are typically circumscribed, reddened mucosal areas with small vessels radiating towards a central scar and are seen in about 5 to 10 percent of patients with IC/BPS.^4^ Diagnosis is made via cystoscopy and confirmed by biopsy. Patients with IC/BPS-HL tend to have more severe symptoms than patients with Interstitial Cystitis/Bladder Pain Syndrome without Hunners Lesions (IC/BPS-NHL).

Evidence for immunologic dysfunction in IC/BPS includes the observation that several autoimmune disorders are significantly more prevalent in patients with IC/BPS than in the general population. These disorders include: Sjögren’s syndrome, which has been found in 7.2% of patients with IC/BPS as opposed 0.5% in the overall population^5^; rheumatoid arthritis, which has been observed in 4-13% of patients with IC/BPS compared to 1.0% of the general population^6^; and systemic lupus erythematosus, which is observed in 1.7% of patients with IC/BPS, as opposed to 0.05% in the general population.^7^

Immune system involvement in IC/BPS is also supported by the elevated counts of immune cells found in the bladder of patients with IC/BPS. Mast cells, among other immune cell types, have been reported to be elevated in the submucosal and detrusor layers of the bladder wall of patients with IC/BPS-HL and IC/BPS-NHL.^8^ Mast cells can produce and release numerous pro inflammatory mediators including Interleukin-6 (IL-6), Interleukin-8 (IL-8), prostaglandins, Histamine, and Vascular Endothelial Growth Factor (VEG-F).^9^ Previous studies have shown elevated levels of IL-6 and IL-8 in the urine of patients with IC/BPS, and have also linked urinary VEG-F levels with higher rates of IC/BPS bladder glomerulations.^10, 11^

Cells found within the bladder biopsies of patients with IC/BPS-HL have also been shown to exhibit expression of several B and T cell markers including cytotoxic T lymphocyte-associated protein (CTLA)-4, Cluster of Differentiation (CD) 20, and CD79A.^12^ Other studies have also confirmed significant elevation in CD138 positive (CD138+) plasma cell counts within the bladder of patients with IC/BPS-HL. Substantial plasma cell inflammation (200+ cells/mm^2^) was observed in 93% of IC/BPS-HL samples and 8 % of IC/BPS-NHL samples.^13^

Another connection between IC/BPS and the immune system is the involvement of the Nerve Growth Factor (NGF) neurotrophin, which has been implicated in IC/BPS pathogenesis.^14^ Multiple immune cells such as Mast cells and Macrophages can induce the increased production of NGF and its concentrations are elevated in the bladder wall of patients with IC/BPS compared to unaffected controls.^15, 16^ Urinary NGF levels, in turn, correlate with urinary urgency and pain severity among patients with IC/BPS, providing a potential mechanistic link between immune cells and disease symptoms.^14^

The majority of studies in the current literature have focused either on soft tissue or urine analysis from generalized IC/BPS patient cohorts and have identified infiltrating immune cells in the bladder, as well as elevated concentrations of multiple cytokines and antibodies in the urine.^17 1, 18, 19^ However, different studies have identified different cytokines as being upregulated in IC/BPS. ^10, 16, 20^ It is possible that variation in the conclusions from these studies is due to inconsistent disease classifications between treatment centers. In addition, while immune cell infiltrates, including C20+ and CD138+ B cells have been observed, a profile of the relative abundance of the different immune cell types within patients is still necessary to clarify the role of the immune system as it pertains to this disease. ^12, 13^ Our study assessed both urinalysis and bladder histology to create bladder immune cell profiles of patients with IC/BPS-HL and to detect differences in urinary cytokine concentrations between IC/BPS-HL, IC/BPS-NHL, and unaffected control (UC) sub-populations. Average numbers of CD138+ and CD20+ B cells, and CD3+ T cells in IC/BPS-HL bladder walls were significantly elevated compared to unaffected controls (UC). Patients with IC/BPH-HL had significantly elevated urinary levels of Interleukin 6 and Tumor Necrosis Factor-α compared to patients with IC/BPS-NHL and UCs.

## Materials and Methods

### Immunohistochemistry staining

All work received prior approval from the Institutional Review Board at Northwell Health. The presence of HL was confirmed by cystoscopy prior to sample retrieval. Thin 4-6 µm tissue sections were cut and stained with Hematoxylin and Eosin (H&E). Using similar thin sections, immunohistochemical (IUC) stains were performed on each biopsy sample for CD3, CD20, CD14, CD15, CD56, and CD138, for the quantification of T cells, B cells, monocytes, eosinophils/neutrophils, Natural Killer (NK) cells, and plasma B cells respectively (**Figure 2A**). Hematoxylin was used as the counterstain. All antibodies were obtained from Ventana medical systems, Tucson, AZ. Antibody product details are as follows: CD3 (Catalog #: 790-4341, Lot F21480), CD20 (760-2531, Lot F18651) CD14 (760-4523, Lot V0001577) CD15 (760-2504, Lot F16036) CD56 (790-4465, Lot F13782) CD138 (760-4248, Lot 20065709).

**Figure 2:**
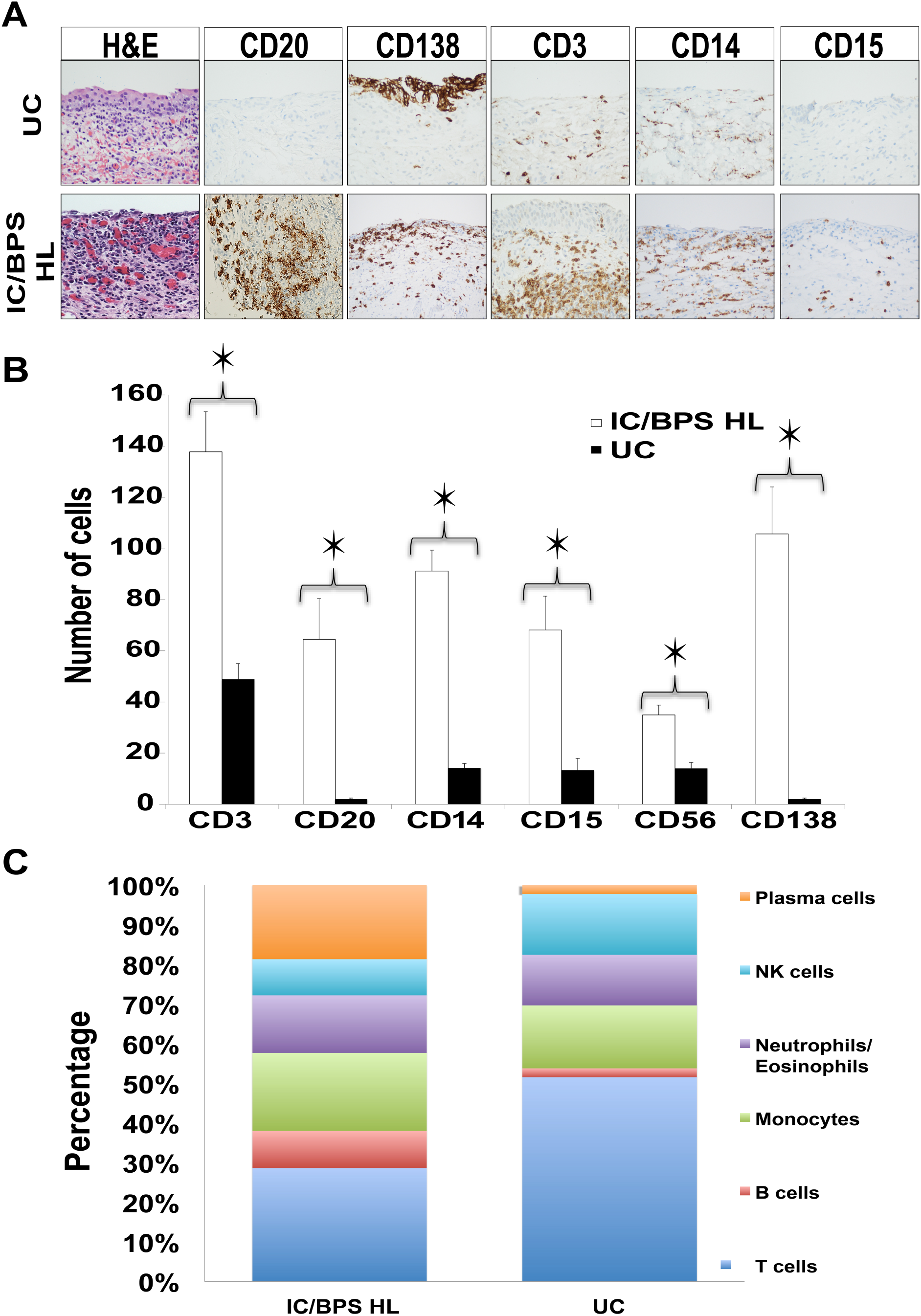
Summary of IUC results. **Figure 2A. Images of biopsied tissues from patients with Interstitial Cystitis/Bladder Pain Syndrome with Hunners Lesions (IC/BPS-HL) and unaffected controls (UC) stained for immune cell markers**. Cross sections of bladder urothelium viewed at 40X magnification and stained for Hematoxylin and Eosin (H&E), (nucleus/cytoplasm) Cluster of differentiation-3 (CD3), (pan T cells), CD20, (B cells) CD138, (plasma cells) CD14 (monocytes), and CD15 (neutrophils/eosinophils). Biopsies were performed on 48 patients diagnosed with IC/BPS-HL and 2 unaffected controls (UC). Thin (4-6 µm) cross sections of bladder tissue were then stained with either H&E or specific CD markers. Images of Hunner’s lesions and control tissue are shown. **Figure 2B. Average cell marker counts found in the bladder biopsy samples of patients with IC/BPS-HL vs. UC**. Cells positive for each marker were quantified per high-dry field (x400), taken in the area of maximum diffuse infiltration by the inflammatory cells. Counts for IC/BPS-HL samples are indicated by white bars, and unaffected controls (UC) by black bars. Error bars indicate standard error of the mean. The differences in observed counts between the IC/BPS-HL and UC cohort were significant at 0.0005 level for all 6 cell markers measured in this study. On average, 498 cells were found in each of the IC/BPS-HL samples compared to only 95 in the control samples (P = 4.17E-11). Average cell counts for every marker were at least 2.5 fold higher in IC/BPS-HL samples versus the unaffected controls. The most drastic differences can be observed in average CD138 counts (IC/BPS-HL N= 106, UC N =2, P value= 1.34E-06) and CD20 counts (IC/BPS-HL N= 65, UC N =2, P value= 3.26E-04). **Figure 2C. Relative proportions of cells displaying each cell marker in UC and IC/BPS-HL cohorts**. The proportion of Cluster of Differentiation (CD)-20 positive (CD20+) B cells (IC/BPS-HL: 9%, UC: 2%) and CD138+ plasma cells (IC/BPS-HL: 19%, UC: 2%) was much greater in patients with IC/BPS-HL compared to the unaffected controls, while the percentage of Natural Killer cells (IC/BPS-HL: 9%, UC: 15%), Monocytes (IC/BPS-HL: 20%, UC: 16%), and Neutrophils (IC/BPS-HL: 14%, UC: 13%) did not differ significantly between the two cohorts. The relative abundance of cells displaying the CD3 T cell marker fell from 52% among the unaffected controls to 29% among the IC/BPS-HL cohort.

Based on the IUC staining, counting was performed and recorded per high-dry field (x400) in the area of maximum diffuse infiltration by the inflammatory cells. Areas of distinct lymphoid follicle or tight aggregate formation were not counted to prevent skewing of results. However, the presence of such lymphoid-predominant areas was noted. Biopsies from negative controls were examined and quantification of cells was performed along with the study cohort. Both staining and cell counting was conducted in a blinded manner. Multiple counts were recorded from each patient and the average cell count for each marker was used during data analysis. A 2-tailed independent samples T-test was utilized to assess the statistical significance of the results.

### Mesoscale Discovery Cytokine assay

Levels of 9 different immunoregulatory cytokines (Interferon (IFN)-γ, IL-1B, IL-2, IL-4, IL-6, IL-8. IL12P70, IL-13, and Tumor Necrosis Factor (TNF)-α) were assessed using a Mesoscale Discovery (MSD) U-plex 10 spot multiplex assay (Mesoscale discovery, Catalog # K15049K-1). After preparing the 96-well plate as per manufacturer instructions, normalized urine samples from 18 IC/BPS-HL, 18 age and sex matched IC/BPS-NHL counterparts, and 4 UC research participants were tested in duplicate. Plate readings were conducted using an MSD Quickplex SQ 120 imager (Model no: 1250) and the data used for analysis was obtained using the MSD workbench software program (version 3.0.18). A 2-tailed independent samples T-test was utilized to assess the statistical significance of the results.

### Creatinine Normalization

Urine from each patient was tested for Creatinine concentration in order to determine the appropriate volumes to be used in the aforementioned multiplex cytokine assay. Samples were run in duplicate and averaged to obtain the final result. An Invitrogen Creatinine urinary detection kit (Thermofisher catalog number: EIACUN) was used and the manufacturer protocol was followed.

## Results

### Sample Collection

Biopsy samples were acquired from the Hunner’s Lesions of 48 patients with IC/BPS-HL after obtaining informed consent. The presence of HL (similar to those seen in **Figure 1**) was confirmed by cystoscopy prior to sample retrieval. Biopsy samples from 2 patients with no morphologic or clinical features of IC or any other urological malignancy were also retrieved, stained, and used as unaffected controls (UC).

Urine samples were obtained from 18 patients with IC/BPS-HL, 18 with IC/BPS-NHL and 4 UC. Urine was first centrifuged to remove heavy debris and then normalized using a creatinine detection kit. Concentrations of urinary creatinine ranged from 11-279 mg/dL.

### Immunohistochemistry

Immunohistochemistry (IUC) is an effective tool to determine the distribution of cell markers of interest within tissue samples.^21^ Previous studies have indicated that there is an increase in immune cells within IC/BPS bladder lesions; the presence of cells displaying B and T cell markers were confirmed by IUC and RT-PCR.^8, 12^ Yet, this previous work did not confirm whether a particular population of immune cells was unusually elevated, as only a few markers were used per study. In the present experiments, a variety of immune cell markers were examined in HL biopsies using IUC.

IUC stains were performed on each of the IC/BPS biopsy samples and UC samples. Markers were selected to reflect the diversity of immune cells. These markers included: CD3 (T cells), CD20 (B cells), CD14 (monocytes/macrophages), CD15 (granulocytes including neutrophils and eosinophils), CD56 (natural killer cells), and CD138 (plasma cells) (**Figure 2A)**. Based on the IUC staining, cells were counted and the number of positive cells was recorded per high-dry field (x400) in the area of maximum diffuse infiltration by the inflammatory cells. Areas of distinct lymphoid follicle or tight aggregate formation were not counted to prevent skewing of results. Statistical significance was assessed using a 2-tailed independent samples T-test.

On average, 498 cells expressing one of the immune cell markers were found in each of the IC/BPS-HL bladder samples compared to only 95 in the control samples (p value = 4.17E- 11). Average counts for cells displaying each individual marker of interest were significantly higher among patients with IC/BPS-HL vs UC’s (**Figure 2B and C**). Strikingly, CD138+ plasma cell counts displayed the largest discrepancies: a more than 50-fold increase in IC/BPS-HL samples (on average,106 cells/field) as opposed to the UC cohort (2 cells/field, p-value: 1.34E- 06 **Figure 2B**). CD20+ B-cell counts were also elevated among the patients with IC/BPS-HL (65 cells/field) when compared to UC (2 cells/field p-value: 3.26E-04).

Other immune cell types were also observed to be more prevalent in the IC/BPS-HL samples than in controls. For T cells, the number of CD3+ cells was significantly elevated in patients with IC/BPS-HL (138/field) as opposed to UC (49 cells/field, p-value: 2.52E-06 **Figure 2B**). Numbers of monocytes and macrophages, as reflected by CD14+ cells were also significantly elevated among the IC/BPS-HL cohort (91 cells/field) vs. the UC group (14 cells/field, p value: 5.24E-12 **Figure 2B**). Likewise, granulocytes including eosinophils and neutrophils, as reflected by CD15+ cell counts were more prevalent in samples from patients with IC/BPS-HL (68 cells/field) compared to UC (13 cells/field, p-value: 2.66E-04 **Figure 2B**). Similarly, patients with IC/BPS-HL showed higher levels of CD56+ NK cells (35 cells/field **Figure 2B**) vs. UC (14 cells/field, p-value: 2.15E-05 **Figure 2B**).

### Quantifications of Cytokines

Previous studies have indicated that patients with IC/BPS have elevated urinary concentrations of various cytokines including IL-6, IL-17, and IL-33.^18, 22, 23^ Cytokines are secreted by cells of the immune system and function as primary mediators and modulators of the immune response.^24^ Therefore, the cytokines observed in IC/BPS patients can provide insights about immune involvement in the disease.

Urine cytokines in the present study were detected using the mesoscale discovery (MSD) U-plex assay: a highly sensitive electrochemiluminescence plate assay. Nine cytokines were selected for testing based on previous literature and role in the immune system: IFN-γ, TNF-α, IL-1β, IL-2, IL-4, IL-6, IL-8, IL-12p70, and IL-13.

IL-6 is a key cytokine involved in B cell activation^25^ and the results from the cytokine assay indicated that patients with IC/BPS-HL, when compared to their age and sex matched IC/BPS-NHL counterparts, displayed significantly higher average urinary concentrations of IL-6 (**Figure 3A**. HL: 3.23 pg/mL, NHL: 1.61 pg/mL, P value = 0.0175). A significant difference in average urinary IL-6 concentrations was also observed between the IC/BPS- HL population and UC population (**Figure 3A**. HL: 3.23 pg/mL, UC: 1.14 pg/mL, P value = 0.0028).

**Figure 3.**
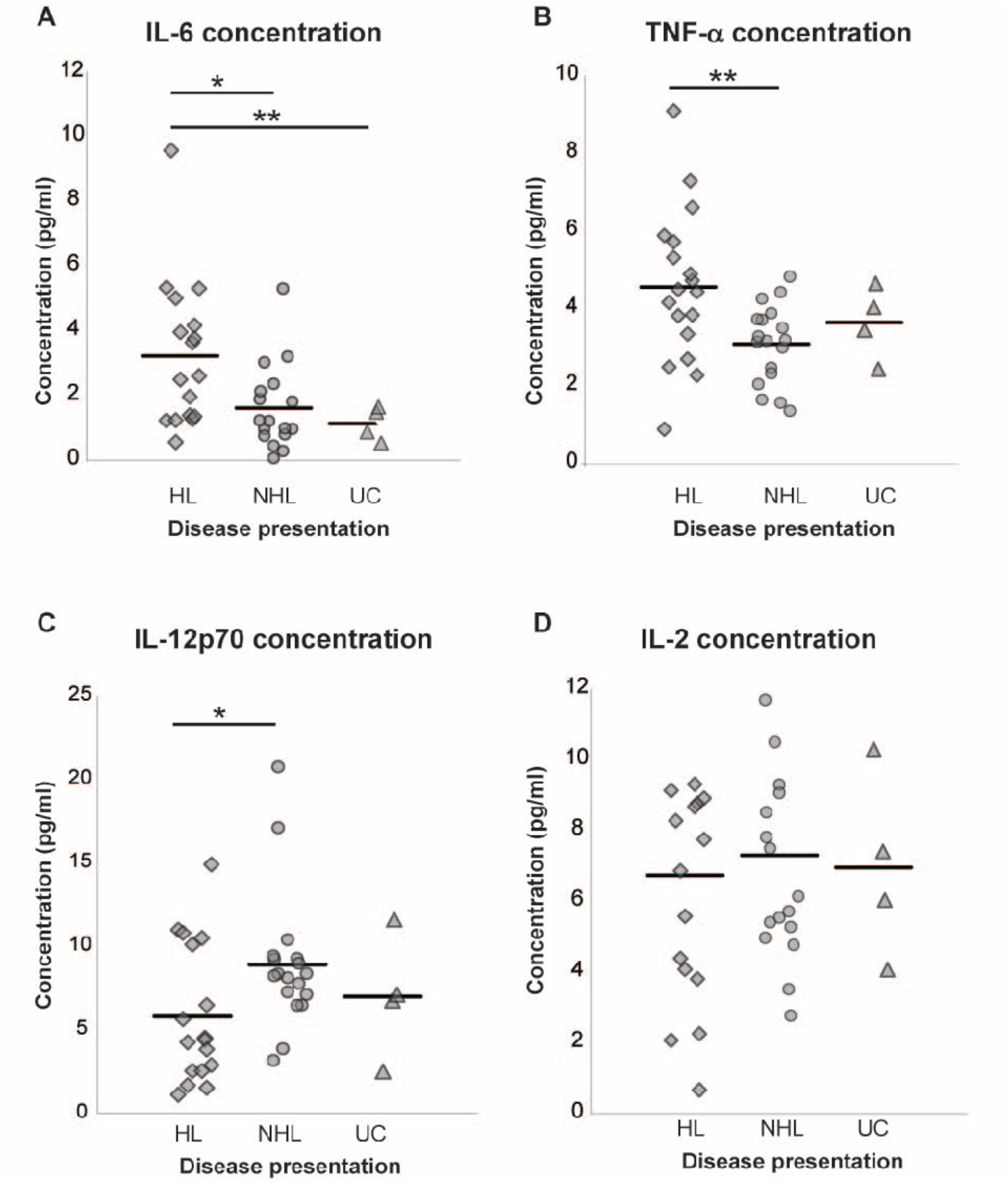
Urinary Cytokine concentrations in patients with Interstitial Cystitis/Bladder Pain Syndrome (IC/BPS), and unaffected controls (UC). Nine cytokines (Interferon-γ, Interleukin (IL)-1β, IL-2, IL-4, IL-6, IL-8, IL-12p70, IL-13, and Tumor Necrosis Factor-α) were assessed using a MSD U-plex 10 spot assay (Mesoscale discovery, Catalog # K15049K-1). Individual and average (indicated by the horizontal bar) cytokine concentrations for **(A)** IL-6, **(B)** TNF-α, **(C)** IL-12p70, and **(D)** IL-2 from patients with Interstitial Cystitis/Bladder Pain Syndrome without Hunners Lesions (IC/BPS-NHL), patients with Interstitial Cystitis/Bladder Pain Syndrome without Hunners Lesions (IC/BPS-HL), and UC research participants. Patients with IC/BPS-HL, compared against their age and sex matched IC/BPS-NHL counterparts, displayed significantly higher average urinary concentrations of IL-6 (HL: 3.23 pg/mL, NHL: 1.61 pg/mL, P value = 0.0175) and TNF-α (HL: 4.55 pg/mL NHL: 3.08 pg/mL, P value = 0.009). A significant difference in average urinary IL-6 concentrations was also observed between the IC/BPS- HL population and unaffected control (UC) population as well (HL: 3.23 pg/mL, UC: 1.14 pg/mL, P value = 0.0028). The IC/BPS-HL cohort also displayed significantly lower urinary concentrations of IL-12p70 compared to the IC/BPS-NHL cohort (HL: 5.81 pg/mL, NHL: 8.91 pg/mL, P value = 0.0334). No other statistically significant differences in urine cytokine concentrations were observed among the 3 sub-groups. Urine from 18 IC/BPS-HL, 18 age- and sex-matched IC-BPS-NHL counterparts, and 4 control (without any inflammatory urological disease) research participants were tested in duplicate. The volume of urine used for each research subject was based upon the results of a prior Creatinine normalization assay. The data used for analysis was obtained using the MSD workbench software program (version 3.0.18).

TNF-α serves as a central mediator of the immune response and is produced by numerous immune cells such as mast cells, macrophages, and fibroblasts.^26^ Urinary TNF-α concentrations were also significantly elevated among the IC/BPS –HL cohort compared to the IC/BPS-NHL group (**Figure 3B**. HL: 4.55 pg/mL NHL: 3.08 pg/mL, P value = 0.009).

IL-12p70 is a pro-inflammatory cytokine that plays a key role in inducing T helper 1 (Th1) and Th17 T cells.^27, 28^ Urinary IL-12p70 concentrations were significantly lower in the IC/BPS-HL population compared to the IC/BPS-NHL cohort (**Figure 3C**, HL: 5.81 pg/mL, NHL: 8.91 pg/mL, p value: 0.033). There was no significant difference between UC and HL urinary IL-12p70 concentrations (**Figure 3C**. HL: 5.81 pg/mL, UC: 7.00 pg/mL, p value: 0.60). Urinary concentrations of IFN-γ, IL-1B, IL-2 (**Figure 3D**), IL-4, IL-8, and IL-13 did not yield statistically significant results in our patient cohorts.

## Discussion

This study indicates that there are many significant differences in both the number of immune cells within the bladder and cytokine concentrations in the urine between patients with IC/BPS-HL and unaffected controls.

One of the more striking differences in patients with IC/BPS-HL was the significant elevation in numbers of cells expressing the B lineage CD20 and CD138 cell markers **(Figure 2A and B)**. This drastic increase in the proportion of CD20+ and CD138+ cells indicates that B and plasma cells may have a specific role to play in the development or pathology of Hunner’s lesions. This observation can serve as a starting point for identifying candidate mechanisms, diagnostic markers, or therapeutics for IC/BPS-HL.^19^ These results are consistent with previous findings that also observed significantly increased CD20+ and CD138+ cell counts within the bladders of patients with IC/BPS-HL. ^12, 13, 19^ These previous studies typically analyzed 2-3 markers. The large number of immune cell markers analyzed in the present study allowed for the comparison of multiple cell types and therefore established that the relative increase in the number of B cells is especially noteworthy.

The results from the MSD cytokine assay are also concordant with the observation of increased numbers of B cells in patients with IC/BPS-HL. IL-6 is a key cytokine involved in B cell activation.^25^ Urinary IL-6 concentrations were significantly higher in patients with IC/BPS- HL compared to patients with IC/BPS-NHL (**Figure 3A**). These results suggest that suggesting that IL-6 may play a role in the observed increased numbers of CD20+ and CD138+ cells in the bladder of patients with HL. The results from this MSD assay replicate results from numerous previous studies that also found that urinary IL-6 levels were elevated in patients with IC/BPS- HL.^10, 22^ A prior study in elderly patients confined to a nursing home found a similar level of elevation in asymptomatic bacteriuria. 29 While IL-6 elevation is not specific for IC/BPS-HL, and the small difference between UC and patients with IC/BPH-HL may make the design of a clinical test difficult, our observation of increased IL-6 levels is consistent with B cell infiltration into the bladder and provides an important piece of evidence about the immunology of the disease.

In addition to B cells, the IUC results indicate that, when compared to UCs, patients with IC/BPS-HL displayed significantly higher numbers of CD3+ T cells within their bladders (**Figure 2B**, HL: 138, UC: 49, p value: 2.52E-6). Interestingly, urinary IL-12p70 concentrations were significantly lower among the IC/BPS-HL sub population compared to the IC/BPS-NHL cohort (**Figure 3C**, HL: 5.81 pg/mL, NHL: 8.91 pg/mL, p value: 0.033). IL-12p70 is a pro-inflammatory cytokine that plays a key role in inducing Th1 and Th17 T cells.^27, 28^ There was no significant difference between UC and HL urinary IL-12p70 (**Figure 3C** HL: 5.81 pg/mL, UC: 7.00 pg/mL, p value: 0.60). Additional studies will be necessary to determine if the elevation of IL-12p70 is consistently present in patients with IC/BPS-NHL. Given the relationship between IL-12p70 and T cell activation pathways, further testing could focus on confirming the specific T cell subsets that are upregulated among patients with IC/BPS-NHL and -HL. For instance, further analysis of the types of immune cells involved in IC/BPS-NHL could help determine if these patients have greater activation of Th1 and Th17 cells than patients with IC/BPS-HL.

Significant differences between patients with the IC/BPS-HL and IC/BPS-NHL cohorts were also observed in urinary TNF-α levels (**Figure 3B**, HL: 4.551 pg/mL, NHL: 3.084 pg/mL, p value: 0.009). TNF-α is produced by numerous immune cells such as mast cells, monocytes and macrophages and is a central mediator of the immune response.^26^ The increased urinary TNF-α concentrations found amongst the IC/BPS-HL cohort builds upon previous findings which describe significantly elevated levels of TNF-α in the sera of patients with IC/BPS and identify the cytokine as a possible mediator of bladder urothelium apoptosis, which is observed in IC/BPS.^30, 31^ As TNF-α is increased in overactive bladder as well as IC/BPS,^26^ measurements of TNF-α concentration on their own will be unlikely to produce a stand-alone clinical test for IC/BPS. However, the consistent observation of increased TNF-α in patients with IC/BPS will be useful for establishing immunological models that could ultimately lead to improvements in diagnosis and treatment.

While we used an expanded panel of immune markers, the relative increase in B cells and plasma cells in IC/BPS patients with Hunner’s lesions observed in this study are consistent with prior findings in the field.^13, 19^ Continued analysis of cell populations from IC/BPS patient samples and UC samples will help support this emerging consensus on the role of B cells in IC/BPS, while more mechanistic experiments, potentially with animal models, could help test aspects of their contribution to the disease.

Given the increased proportion of plasma cells found in the bladder of patients with IC/BPS and strong association between Toll-Like Receptor (TLR) activity and IC/BPS severity, ^32^ it is possible that TLR activity mediated through a B cell activation pathway is involved in IC/BPS pathogenesis. TLR’s are involved in T cell independent B cell activation, and increased TLR-4 signaling has been associated with increased pain and symptom severity among women with IC/BPS, and other inflammatory pain conditions, including rheumatoid arthritis.^32, 33-36^ In patients with IC/BPS, elevated TLR activity correlates with increased pain frequency, pain intensity, higher rates of reduced sexual functioning, and overall higher Genitourinary Pain Index scores.^32^ *In vitro* stimulation of B cells by TLR’s has been shown to induce B cell proliferation and differentiation into activated antibody-producing plasma cells and *in vivo* TLR signaling contributes to T cell independent antibody responses to bacterial infection.^37^ Thus, it is possible that the increased numbers of B cells and plasma cells we detected in the bladder of IC/BPS patients is due to the activation of TLR-4.

## Conclusion

Our results indicate that activation of B-cells and plasma cells correlates with IC/BPS-HL disease status. This conclusion is based on the high number of CD138+ and CD20+ cells found in the bladder tissue and the significant increase in IL-6 urinary concentrations among the HL cohort. While the present study has identified several immune cells present in the HL, the study design cannot determine whether the presence of these cells is a consequence or a cause of disease. It is likely that the increase in immune cell counts and coinciding upregulation of certain pro inflammatory cytokines represent a disease process in IC/BPS, but we cannot exclude that their presence may be neutral or even protective. Future studies should address the relationship between immune cell activation and IC/BPS disease severity in order to establish a causal relationship.

We did not find a statistically significant correlation between the concentration of cytokines in the urine of patients and the severity of clinical symptoms such as urinary urgency or severity of pain. The interpretation of this result is complicated because we cannot be certain whether it is due to natural variability in the levels of cytokines between different people or a true lack of a relationship between cytokine levels and disease. Longitudinal analysis of the relationship between clinical symptoms and degree of inflammatory changes can help resolve this issue.

It is still unclear how chemokines and cytokines in urine or serum correlate to bladder immune infiltration. Since chemokines and cytokines are indicators of inflammation, exploring the relationship between them and bladder immune cells in IC/BPS should be a high priority for future research. In the present study, biopsy samples were collected asynchronously with urine samples, so a correlation analysis could not be performed. While it is not always technically or ethically possible to collect urine and biopsy specimens from a given patient simultaneously, it is a direction that should strongly be considered in future studies.

## Data Availability

As per IRB guidelines, all clinical data pertaining to this study are stored within secure Northwell Health cloud based storage systems and all hard copy documentation is stored within access controlled Northwell Health facilities. Data can be made available upon request.

## Acknowledgements

The research team would like to acknowledge Jeffers Nguyen for assisting with cytokine selection methodology for the MSD assay and Billal Ahmed for his time spent compiling data and assisting with bench-top assays.

## Funding

I.T. was supported by a pilot grant from the Kavli Foundation. V.N., M.F., W.D., and J.N.H.S. were supported by a grant from the H.F. Langbert Neuroimmunology Research Fund.

## Supplemental Material

**Supplemental Table 1:**
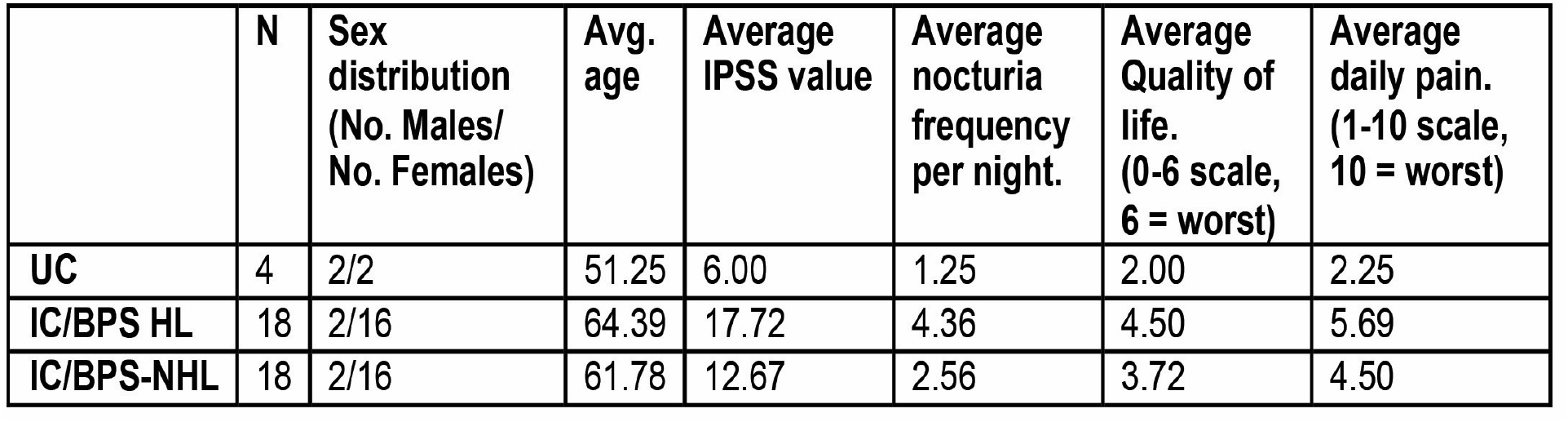
Clinical characteristics and sex breakdown of the 3 research cohorts whose urine was collected for analysis with the Mesoscale discovery (MSD) multiplex cytokine assay. Proper informed consent was obtained from all patients prior to sample retrieval. (IPSS: International Prostate Symptom Score; UC: Unaffected controls; NHL: Patients with Interstitial Cystitis/Bladder Pain Syndrome without Hunner’s Lesions; HL: Patients with Interstitial Cystitis/Bladder Pain Syndrome with Hunner’s Lesions).

|  | N | Sex distribution<br>(No. Males/<br>No. Females) | Avg.<br>age | Average<br>IPSS value | Average<br>nocturia<br>frequency<br>per night. | Average<br>Quality of<br>life.<br>(0-6 scale,<br>6 = worst) | Average<br>daily pain.<br>(1-10 scale,<br>10 = worst) |
| --- | --- | --- | --- | --- | --- | --- | --- |
| UC | 4 | 2/2 | 51.25 | 6.00 | 1.25 | 2.00 | 2.25 |
| IC/BPS HL | 18 | 2/16 | 64.39 | 17.72 | 4.36 | 4.50 | 5.69 |
| IC/BPS-NHL | 18 | 2/16 | 61.78 | 12.67 | 2.56 | 3.72 | 4.50 |

**Supplemental Table 2:** Clinical characteristics and sex breakdown of the 2 research cohorts who donated biopsies for immunohistochemistry analysis of bladder tissue samples. Proper informed consent was obtained from all patients prior to sample retrieval. Since UC patients did not present with urological symptoms, they were not asked about clinical information regarding their voiding and urinary associated pain, and therefor this information was not available in their clinical records. (UC: Unaffected controls; IC/BPS-HL: Patients with Interstitial Cystitis/Bladder Pain Syndrome with Hunner’s Lesions)

|  | N | Sex distribution<br>(No. Males/<br>No. Females) | Avg.<br>age | Average<br>nocturia<br>frequency<br>per night. | Average<br>Quality of<br>life.<br>(0-6 scale,<br>6 = worst) | Average<br>daily pain.<br>(1-10 scale,<br>10 = worst) |
| --- | --- | --- | --- | --- | --- | --- |
| IC/BPS-HL | 48 | 10/38 | 61.6 | 4.53 | 4.81 | 6.26 |
| UC | 2 | 2/0 | 68.5 | n/a | n/a | n/a |

## Notes

### Competing Interest Statement

The authors have declared no competing interest.

### Funding Statement

This project was supported by grants received from the Kavli Foundation and H.F. Langbert Neuroimmunology Research Fund.

### Author Declarations

All research protocols and study related documents were assessed and approved by the Institutional Review Board at Northwell Health (Study number 17-0254 NSUH).

